# WikiProject Clinical Trials for Wikidata

**DOI:** 10.1101/2022.04.01.22273328

**Authors:** Lane Rasberry, Sheri Tibbs, William Hoos, Amy Westermann, Jeffrey Keefer, Steven James Baskauf, Clifford Anderson, Philip Walker, Cherrie Kwok, Daniel Mietchen

**Affiliations:** University of Virginia; Duke University; ClinWiki; New York University; Vanderbilt University; University of Virginia and Ronin Institute

**Keywords:** ClinicalTrials.gov, Wikidata, Wikipedia, multilingual information, content translation, clinical trials, FAIR data, open data

## Abstract

WikiProject Clinical Trials is a Wikidata community project to integrate clinical trials metadata with the Wikipedia ecosystem. Using Wikidata methods for data modeling, import, querying, curating, and profiling, the project brought ClinicalTrials.gov records into Wikidata and enriched them. The motivation for the project was gaining the benefits of hosting in Wikidata, which include distribution to new audiences and staging the content for the Wikimedia editor community to develop it further. Project pages present options for engaging with the content in the Wikidata environment. Example applications include generation of web-based profiles of clinical trials by medical condition, research intervention, research site, principal investigator, and funder.

The project’s curation workflows including entity disambiguation and language translation could be expanded when there is a need to make subsets of clinical trial information more accessible to a given community. This project’s methods could be adapted for other clinical trial registries, or as a model for using Wikidata to enrich other metadata collections.

## Introduction

WikiProject Clinical Trials is a project to integrate clinical trial metadata into the Wikipedia platform. The motivation for doing so is to increase access to information about clinical trials, to encourage Wikipedia-style public and multilingual curation of the content, and to facilitate discussion and understanding of the research metadata. The project imported ClinicalTrials.gov records into Wikidata, which is a structured data complement to Wikipedia.[1] Once those records were in Wikidata, the project enriched the original content using content curation workflows which are common in that community, and which we present as a model of engagement for sharing data in Wikidata. As model outcomes of this process, we show Wikidata profiles of clinical trials for a medical condition, an intervention, a research site, a principal investigator, and a funder. Complementary to this, we translated some clinical research terms in Wikidata to showcase Wikidata’s multilingual capacity to present profiles in multiple languages. Users can access this content in any way that they access Wikidata, but as a portal and starting point, WikiProject Clinical Trials presents a browsing interface and links to resources.[2] Snapshots of the website and model data outcomes are available in a February 2022 archival record.[3] Figure 1 shows the project landing page where links to available resources include information about the data model, query collection, instructions to curate, details about the project, and forum to discuss the project.

**Figure 1.**
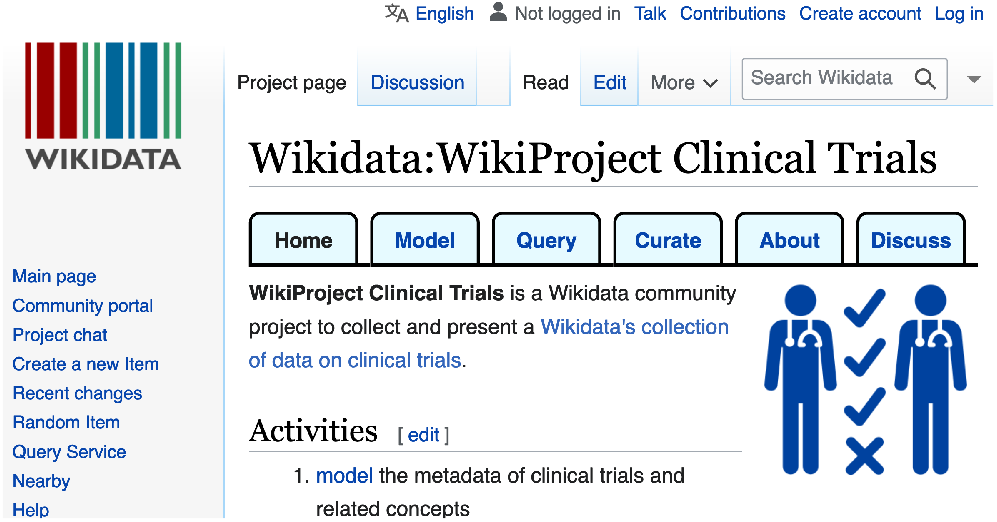
[Wikidata WikiProject Clinical Trials screen-shot](https://commons.wikimedia.org/wiki/File:Wikidata_WikiProject_Clinical_Trials_screenshot_2022-02.png). In the Wikimedia ecosystem, “WikiProjects” serve as virtual community forums for inviting public participation, sharing resources, and hosting discussions. WikiProject Clinical Trials shown here links to resources labeled “Model”, “Query”, “Curate”, “About”, and “Discuss”.

In this paper, we use various “wiki” terms, including Wikipedia, Wikimedia, and Wikidata. To clarify, all of these names refer to different aspects of a single platform which includes a virtual environment, community, media collection, and culture. “Wikipedia” is the name of the encyclopedia and its community; “Wikidata” is for the general reference data project; and “Wikimedia” is the term for integrative activity such as combining Wikipedia and Wikidata. This paper uses these terms as the Wikimedia community itself does.

Desired outcomes of this project include circulation of clinical trials metadata on the open web as linked open data and the exploration of using the Wikipedia platform as a portal for access to clinical trials data associated with a given medical condition, treatment, researcher, or organization. We take a position that access to clinical trials metadata should be universal, and show our Wikidata language translation and Wikidata’s multilingual capacity as progress toward better access.

ClinicalTrials.gov is the United States government public clinical trials registry for clinical trial metadata.[4][5] Its metadata includes the name of the trial, the National Clinical Trial number (NCT ID) as an identifier assigned by the registry, the names of key people involved, relevant organizations and their roles such as research sites or sponsors, the treatment intervention examined, and the medical condition targeted. The information is available in English. A readability assessment reported that its trial records were difficult to read.[6] Researchers have expressed wishes that the data in ClinicalTrials.gov were more accessible.[7, 8]

WikiProject Clinical Trials seeks to import ClinicalTrials.gov data into the Wikipedia ecosystem to apply the benefits of the existing Wikimedia infrastructure to clinical trials metadata for the purpose of increasing accessibility, crowdsourced curation, and stakeholder engagement. To be clear, the ClinicalTrials.gov registry and Wikidata only share information that is already public, and WikiProject Clinical Trials is a project to specifically share public metadata about trials in an enhanced fashion.

While the intent of this project is to make data more accessible, at the same time we recognize that our data import and curation process is incomplete as compared to the original dataset. Although the project imported nearly every ClinicalTrials.gov record into Wikidata, only some fields for each record were imported. Wikipedia has a long history as a target for various criticisms, including incompleteness. Similar critiques apply to Wikidata as well. We affirm that incompleteness is undesirable, but also recognize the discourse that Wikipedia’s incomplete publishing has also been part of its development process toward more complete and quality topic coverage.[9]

As Wikipedia belongs to the world’s most popular information sources,[10] it has been the ongoing subject of academic research.[11] A crowdsourced community of editors, most of whom are volunteers, develop its content.[12] Medical information in Wikipedia is popular, and researchers have examined its quality.[13]. On that basis, since the launch of Wikidata, [14] similar research has been undertaken on its quality, [15] including its medical content [16, 17, 18, 19], multilingual aspects [20, 21] and the potential for integration with research workflows.[22, 23, 24, 25, 1]

The Wikimedia editorial community has created and maintains a governance process which uses consensus of participants as the basis of authority.[12] One of the community values is encouraging universal accessibility through language translation of content, so content develops beyond the original language.[26] Wikipedia curates general reference prose while related projects in the Wikimedia ecosystem curate other media types.[27] Various research projects have reviewed and reused Wikipedia as a data source.[28] Wikidata is the project in the Wikimedia ecosystem which presents and curates general reference data.[29] Search engines reuse Wikidata content, making it available to those who use Internet search in many locations.[30] Various commentators have recognized Wikidata as a project with potential to support research, either for research only in Wikidata or as a source of general reference data which integrates with other kinds of information.[22, 31, 32] These characteristics of the Wikimedia platform were factors in our decision to use it for hosting this data.

Collaborating researchers from several institutions each provided useful contributions for developing this project. From Duke University, a contributor imported data from the Aggregate Content of ClinicalTrials.gov (AACT) database, which structures the content of ClinicalTrials.gov to make it more accessible. The nonprofit organization ClinWiki supports patient and research participant communities in curating ClinicalTrials.gov data.[33] ClinWiki shared their data to this project in support of increased patient access. Vanderbilt University has an ongoing effort to increase openness of its research metadata for various benefits, including increased research collaboration and community awareness.[34] As part of that effort, they integrated lists of their own faculty, publications, and clinical research into Wikidata with their Vanderbot project, which makes that university suitable as a test case for researcher profiles. The Patient-Centered Outcomes Research Institute (PCORI) funds research in advocacy of patients.[35] While ClinicalTrials.gov records do not differentiate funding from other sorts of collaboration, our team chose to note PCORI as a funder in Wikidata records. The School of Data Science at the University of Virginia co-develops the WikiCite and Scholia projects, which are efforts to open scholarly publication metadata and use it through Wikidata to create scholarly profiles.[36][37] All of these stakeholders have a common desire to increase access to FAIR and open metadata pertaining to clinical trials.

The name “WikiProject Clinical Trials” refers to the Wikipedia cultural practice of welcoming editors into community collaborations called “WikiProjects”. We established WikiProject Clinical Trials according to community norms which include establishing a project page, posting documentation, and inviting discussion about the content in a public forum for the project. WikiProject Medicine is a documented example of a WikiProject, and our team sought to emulate practices which that project had established.[38, 39]

Our wish as we described it in our original project proposal [40] is that the models we present here support others in leveraging the Wikimedia platform as a hub for new access, content creation, and public understanding in clinical trials data.

## Methods

We divided the development of WikiProject Clinical Trials into a five-step sequence of data modeling, importing, querying, curating, and profiling. As Wikidata is a public platform which invites user engagement, there are points in this process where the community responds to the content we uploaded. Figure 2 presents a model of these steps within the Wikidata editorial process. Branching from our five-step process, the Wikidata reviewers take content which we publish in Wikidata to either edit it or send it onward to any WikiProject for further engagement. Once content is in Wikidata, anyone may use it for the intended use case or any other purpose. The Wikidata community discusses use cases in WikiProject community forums, which are themselves staging grounds for yet more review and the solicitation of off-wiki outreach. Contributions from off-wiki include expert review and further data contributions.

**Figure 2.**
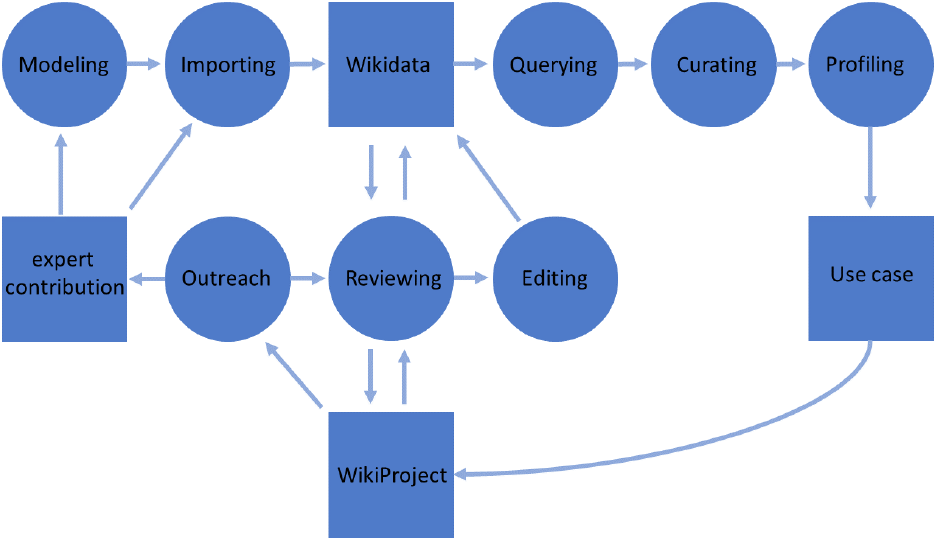
[Wikidata development diagram for WikiProject](https://commons.wikimedia.org/wiki/Wikidata_development_diagram_for_WikiProject.png). WikiProject Clinical Trials landing pused the content curation method visualized here to plan project development. The circles represent actions which we took, and the squares represent points in the process which published records. We describe this process in the methods section..

The model shown here is our own, but we believe that it is suitable as a description for many WikiProjects. Because the design of the Wikidata interface and the steps of its operation change with technological development more quickly than the community’s philosophy of data management, we are sharing this diagram and the below methodology descriptions so that anyone can read Wikidata documentation and consult with the editorial community on how to manage a multi-step project. Technical instructions on how to perform actions become dated quickly, so for current guidance on using Wikidata, the on-wiki help documentation is the best source of information.[41]

### Modeling

Modeling in Wikidata is the process of matching the data for import with corresponding Wikidata properties. The result of this effort was a data model corresponding to the Wikidata item for clinical trial, uniquely identified as Q30612. This model includes a set of Wikidata properties that define basic attributes of a trial. Table 1 presents a set of the properties to which we mapped data from ClinicalTrials.gov. Additional properties, example applications, and links to property documentation for the WikiProject Clinical Trials data model are on the “Model” subpage.[42]

**Table 1.** [WikiProject Clinical Trials data model](https://www.wikidata.org/wiki/Special:PermanentLink/1586085715) (partial). The property label is the English term for the concept and the identifier is the corresponding Wikidata property code. These identities link to Wikidata property pages with more structured data definition and discussion.

| Wikidata property label | identifier |
| --- | --- |
| country | P17 |
| start date | P580 |
| sponsor | P859 |
| number of participants | P1132 |
| title | P1476 |
| short name | P1813 |
| medical condition treated | P2175 |
| ClinicalTrials.gov ID | P3098 |
| research intervention | P4844 |
| clinical trial phase | P6099 |
| research site | P6153 |
| subject recruitment status | P8005 |
| funder | P8324 |
| principal investigator | P8329 |
| study type | P8363 |

Wikidata properties each have a documentation page defining their intent and scope. When anyone imports data to Wikidata, those Wikidata property definitions become labels for the imported data. Our modeling focused on the clinical trial records themselves, and anticipated synergy with other datasets and data models in Wikidata. For example, we had no need to develop models for researchers, organizations, medical conditions, or drugs, because the Wikidata community had already modeled these things and developed precedents for data import. Other projects have also shared methods for importing research into Wikidata, such as for medical resources[43] and for researchers themselves.[44] We could therefore anticipate that by aligning the WikiProject Clinical Trials data model with properties used in other projects, synergies such as links between principal investigators and their institutional affiliations would arise.

Part of the modeling process included the creation of new Wikidata properties through the Wikidata Property Proposal process.[45] In that process, anyone can define new potential properties and propose that Wikidata community reviewers create them following a consensus decision. Example properties which applied to clinical trials and which existed prior to this project include country, start time, sponsor, main subject, number of participants, title, short name, and medical condition treated. New properties which WikiProject Clinical Trials established were research intervention, clinical trial phase, research site, research subject recruitment status, funder, principal investigator, study type, and the ClinicalTrials.gov identifier itself. The team chose these based on opinion that they were of general interest and that they would integrate well with data collections which were already in Wikidata.

### Importing

Once the Clinical Trials Wikidata data model was created, data acquisition and import may proceed by mapping properties in the original dataset to those of corresponding Wikidata property. The original dataset is the clinical trial registry at ClinicalTrials.gov. Instead of taking data from the registry directly, we imported the Aggregate Analysis of ClinicalTrials.gov (AACT) dataset from the Clinical Trials Transformation Initiative because it restructures the original content for increased usability as a dataset.[7] Figure 3 shows the Clinical Trials Transformation Initiative’s model of ClinicalTrials.gov data. WikiProject Clinical Trials data model in Table 1 is only a small part of this complete data schema, and does not attempt complete data mirroring. Because the original dataset was so large, WikiProject Clinical Trials imported only the parts of the original dataset which the team felt could integrate well with Wikidata’s current content holdings and community interests. Reasons for avoiding indiscriminate upload of content include that all uploads trigger the consumption of scarce volunteer labor for review, and that community custom asks that uploaders share content which they reasonably expect others to use.

**Figure 3.**
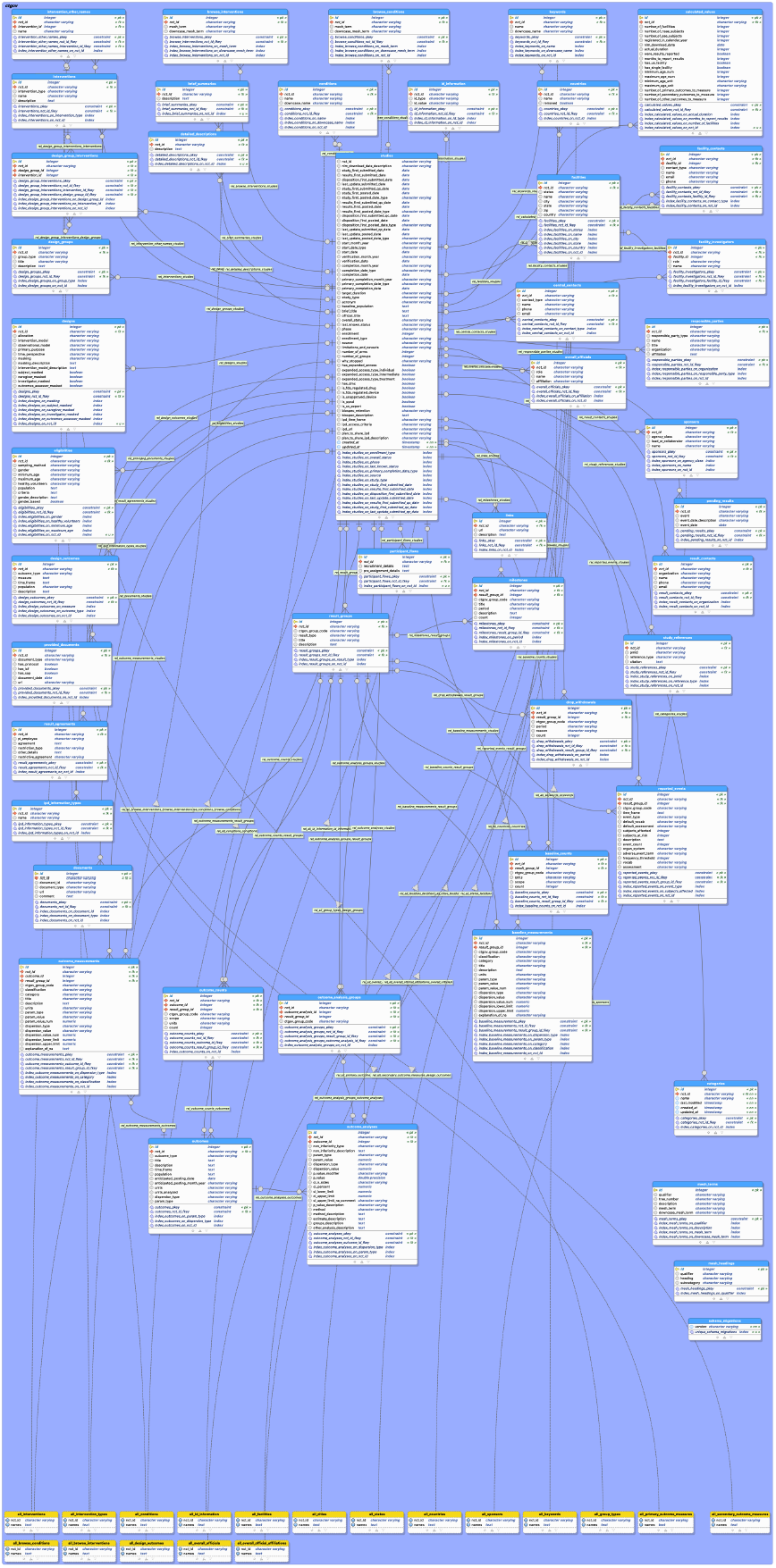
Aggregate Analysis of ClinicalTrials.gov data schema. The Clinical Trials Transformation Initiative develops the Aggregate Analysis of ClinicalTrials.gov (AACT).[48] AACT presents the ClinicalTrials.gov dataset in a way that is easy to access and reuse. Their data schema shown here is more complex than the model which WikiProject Clinical Trials used.

Wikidata redistributes its content holdings with Creative Commons Zero copyright licensing.[46, 47] This means that any data entering the Wikidata environment must become available with that license. Based on Wikimedia community experience in evaluating copyright status, our team determined that the parts of the ClinicalTrials.gov dataset and its AACT adaptation that were identified for import were copyright compatible with Wikidata.

To accomplish data import for this project, one of us (ST) developed and used a tool called the Clinical Trials Wiki Loader. The tool iterates over all clinical trials in the AACT database, mapping attributes to their corresponding Wikidata properties and generating a collection of files containing clinical trial data formatted for ingest via the MediaWiki API QuickStatements tool hosted by the Wikimedia platform’s ToolForge service. Table 4 lists these tools and others, all of which are part of the curation method but some of which contribute to other WikiProject Clinical Trials methods.

Whereas many people are familiar with managing their own data in their own system, Wikidata is a community environment where tens of thousands of editors share virtual space, resources, and volunteer review. Although any user may import datasets into Wikidata, the community in the Wikimedia environment has expectations that content contributors will comply with platform’s social customs.

All uploads to Wikidata are subject to Wikidata community approval through its governance structure, with larger or more integrated uploads getting more scrutiny. Another aspect of Wikidata which may surprise those unaccustomed to co-working a community is that Wikimedia editors always have content reuse on their minds, and may begin repurposing imported content within the Wikimedia platform and elsewhere. Wikimedia governance and community interactions have been described in more detail elsewhere.[49, 12, 50] Before importing content, users should anticipate becoming the subject of attention and be ready to discuss issues publicly if approached. For WikiProject Clinical Trials, the upload process included seeking conversation and review in Wikidata’s public forums before executing data import or large numbers of automated edits. Conversation started with the import of a single data record to confirm its appropriateness. If a reviewer criticized, then the team adapted; if reviewers supported, then the team repeated this cycle with 10 uploads, then 100, 1000, and so on such that the number of participants and depth of review increased with the number of Wikidata edits. The objective in this iterative process is collaborative peer to peer review and friendly social exchange with volunteers. There are no technical criteria for establishing good social relationships, so good Wikidata practice is whatever leads to Wikidata community approval and respect. In the case of WikiProject Clinical Trials, having a project page served to increase transparency and assist reviewers in understanding the project’s intent. The Wikidata review process does not usually create media records for approval, such as is common in social media where “like” buttons are used. Approval should be explicit when there is controversy or some special community request.

### Querying

Data querying in Wikidata is the process for automatically collecting subsets of Wikidata content, typically for curating, profiling or reuse. In human readable terms, WikiProject Clinical Trials queries start with the request “list all instances of (Wikidata property P31) clinical trials (Wikidata item Q30612)”. Next, each of the model queries has a qualification where some additional Wikidata property must match some target values, e.g. particular Wikidata items. Table 2 shows property and example pairs for the project’s five model cases. Queries can use any Wikidata properties and targets in any combination.

**Table 2.** [WikiProject Clinical Trials model profiles](https://www.wikidata.org/w/index.php?oldid=1585305957##Model_profiles). Model profiles are selected to demonstrate results from commonly requested queries.

| Property queried | Example target of query |
| --- | --- |
| main subject (P921) | Zika fever (Q807186) |
| intervention (P4844) | COVID-19 vaccine (Q87719492) |
| research site (P6153) | Vanderbilt University (Q29052) |
| principal investigator (P8329) | Julie McElrath (Q22006776) |
| funder (P8324) | PCORI (Q7144950) |

The Wikidata Query Service is the primary tool in Wikidata for querying to get data and visualizations.[30, 51] WikiProject Clinical Trials presents its queries on a project subpage along with link to run the query through that query service.[52] Table 3 lists the query titles and links to them, and the queries with a February 2022 snapshot of results are archived.[3] The service accepts queries in the SPARQL language (with some added functionality), but even without knowledge to design queries in that language, users may change the search targets by substituting the Wikidata item for one entity with that for another. The current list of queries are on the WikiProject Clinical Trials Query page, while the February 2022 queries and results are archived.[52]

**Table 3.** [WikiProject Clinical Trials Queries](https://www.wikidata.org/wiki/Special:PermanentLink/1585305957). February 2022 results from these queries are archived.[3] Users can change the query targets to engage with Wikidata content as they like.

| Model profiles |
| --- |
| Clinical trials for Zika fever |
| Clinical trials using COVID-19 vaccine |
| Clinical trials at Vanderbilt University |
| Clinical trials with Julie McElrath as principal investigator |
| Clinical trials funded by Patient-Centered Outcomes Research Institute |
| Topics by count of clinical trials |
| Medical conditions |
| Research interventions |
| Research sites |
| Principal investigators |
| Funders |
| Organizational affiliations |
| Clinical trials with principal investigator and their affiliation |
| Clinical trials where principal investigator has Vanderbilt University affiliation |
| Chart of organizations by count of clinical trials |
| Clinical trials where the sponsor was Pfizer |
| Researcher demographics |
| Count of principal investigators by gender |
| Clinical trials where the principal investigator is female |
| Principal investigators by occupation |
| Scope of Wikidata’s clinical trials content |
| List of clinical trials |
| Count of clinical trials |
| Most common properties applied to clinical trials |
| Count of statements in clinical trial records |
| Count of trial records in Wikidata per clinical trial registry |

### Curating

Data curating refers to any of the procedures used to enrich Wikidata content through data disambiguation, correction, or adding missing data. For this project, “curation” often meant interpreting ambiguity in the original data to make it conform to some structured data concept in Wikidata. For example, sometimes the name of a researcher was ambiguous due to being abbreviated or common among various researchers, and the curation was any process which linked them to a corresponding individual as identified by a dedicated Wikidata record. Wikidata is a shared community space, and as such, the Wikimedia community review process oversees all content contributions to maintain and refine its quality standards.[53] Some of the project’s curation methods are described in the curation page of WikiProject Clinical trials.[54] The Wikimedia platform logs the edit history of changes to its content. Commentators have suggested ways to interpret these data logs to understand the community’s editorial process.[55] It is beyond the scope of this project to interpret the intent or outcome of the Wikidata community’s edits to this project’s imported content, but the Wikimedia platform stores the log of edits for each record in a history browsing interface which also includes a suite of Wikimedia tools for interpreting those edits. The majority of trial records imported received an unsolicited edit from a Wikidata user outside the project team.

Table 4 lists software used by WikiProject Clinical Trials with links and persistent identifiers for each. Various workflows in WikiProject Clinical Trials used some software, but the overall data curation process used all of these tools, so presented here are brief descriptions of what each does. The Author Disambiguator assists in matching author name strings of text with personal identifiers. It is configured to identify authors of scholarly publications, and in cases where principal investigators also publish research, this tool helped identify them. The Clinical Trials Wiki Loader is a tool for importing large datasets into Wikidata through the MediaWiki API. ClinWiki is a platform for inviting and managing crowdsourced edits of clinical trial data. Jupyter Notebook is the computational environment that we used for archiving our queries along with the query results. Mix’n’match is a Wikidata tool for matching Wikidata items to identifiers in other databases, and assists with disambiguation. OpenRefine is a tool for matching text strings to Wikidata items. Vanderbot is the Vanderbilt University tool which imports university research metadata to Wikidata. QuickStatements is a Wikidata browser tool for importing medium-sized tabular data to Wikidata. Scholia is a Wikidata-based tool which generates scholarly profiles. TABernacle is a browser-based Wikidata tool which generates editable spreadsheets of Wikidata content based on queries; in this project, we used it to manage language translation. Wikidata is the platform itself. The Wikidata Query Service is the browser based SPARQL interface which is the main route for querying Wikidata. WikiProject Clinical Trials is the web interface which we present for accessing and understanding the data and process of our project.

**Table 4.** Software used by WikiProject Clinical Trials. The software column is the name and link, while RRID and Wikidata items are identifiers assigned by SciCrunch and Wikidata.

| Software | RRID | Wikidata item |
| --- | --- | --- |
| Author Disambiguator | SCR_021247 | Q76693569 |
| Clinical Trials Wiki Loader | SCR_021296 | Q107068605 |
| ClinWiki | SCR_021297 | Q96777166 |
| Jupyter Notebook | SCR_018315 | Q105099901 |
| Mix'n'match | SCR_021302 | Q28054658 |
| OpenRefine | SCR_021305 | Q5583871 |
| VanderBot | SCR_021298 | Q107014192 |
| QuickStatements | SCR_021300 | Q20084080 |
| Scholia | SCR_021248 | Q45340488 |
| TABernacle | SCR_021299 | Q26882268 |
| Wikidata | SCR_018492 | Q2013 |
| Wikidata Query Service | SCR_021301 | Q20950365 |
| WikiProject Clinical Trials | SCR_021311 | Q21829773 |

Since disambiguation was such a focus of this project, we explain “author disambiguation” as an example. Author disambiguation typically refers to the identification of an author of an academic paper, such as when multiple people have the same name, or the author is indicated only by surname and initials. When the principal investigator of a clinical trial also had an academic publication history, the project was sometimes able to disambiguate their Wikidata record to link them to publications, clinical trials, institutional affiliations, and other aspects of scholarly profiling. The Wikidata Author Disambiguator tool assisted with this. The WikiCite project within Wikidata is the broad effort to do metadata curation of this sort.[56] Even when investigators did not have academic publication histories, the principles behind the concept of author disambiguation could still most often identify the person.

Another curation process which we did was human language translation in an effort to increase multilingual access to clinical trials information from the inception of WikiProject Clinical Trials. Wikidata is multilingual and seeks to serve all language communities.[20] One aspect of content curation is connecting or proposing equivalent terms across languages, such that users may query and receive results profiles in any language where structured data exists. Complete translation of medical vocabularies was too complex of an undertaking for this project, so instead, the project’s goal was to set a precedent for ongoing conversation and formation of best practices. ClinicalTrials.gov publishes a “Glossary of Common Site Terms” to explain research to the public.[58] We created Wikidata items for these terms, converted them into simple English, then sent the terms to translation teams for Hindi, Bangla, and Swahili. We chose these languages because they are underrepresented in Internet media, because communities using those languages are research participants in clinical trials, and because those language communities have active participation in Wikidata.

We organized translation with the intent to create cases where users may make queries or receive results using the translated terms, and also to make precedents and documentation for the technical and social process of doing translation of research terms in Wikidata with translators. We accomplished translation in various ways, including by using the Wikidata tool TABernacle. TABernacle is an interface where one may list a set of terms in any one language while prompting for anyone else to translate the terms into another language. In contrast to classical document-centric translations, this data-centric way of translating key terms of a controlled vocabulary allows scalable reuse of translations both within and beyond Wikidata. Some descriptions and links to curation workflows are on the WikiProject Clinical Trials Curate page, while the archived version presents documentation from the time of the project.[54]

### Profiling

A Wikidata-based profile is the result of one or more Wikidata queries presented with context to explain the data to tell a story. A simple Wikidata profile could be a published Wikidata query with a title, so that the query and profile gain the context of someone deeming the results of that query as worthy of sharing or engaging.

More complex profiles may present multiple Wikidata queries with more social context or narration. In the case of WikiProject Clinical Trials, users access profiles through the queries page, and the profiles themselves are the query results with the resources and social context of WikiProject Clinical Trials as a place for engaging with this content. Figure 5 presents “Clinical trials at Vanderbilt University”. The results of this simple query provide a profile of clinical research at an institution, and the social context of the profile is the idea that lists of clinical trials for any institution could be more accessible, e.g. for discussion, for further remixing or other forms of engagement.

**Figure 4.**
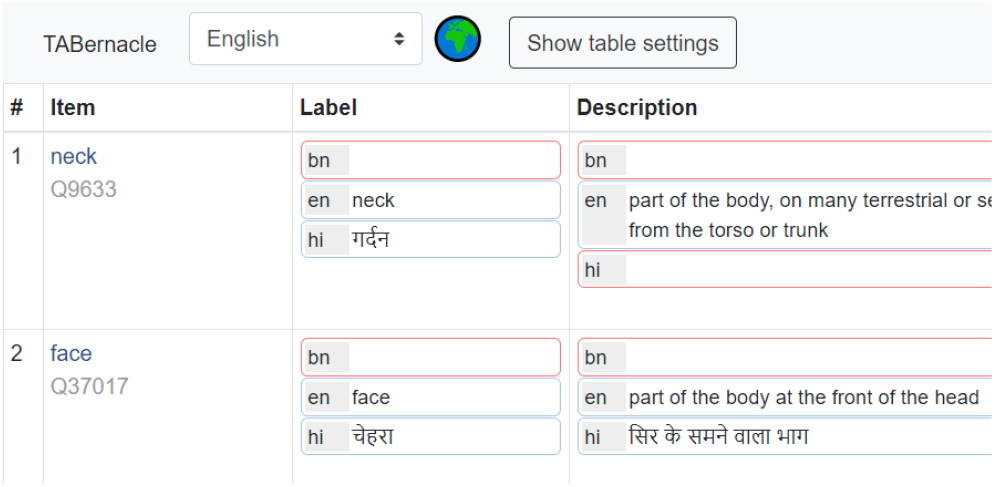
[TABernacle screenshot](https://commons.wikimedia.org/wiki/File:TABernacle_translation_English_Hindi_Bangla.png). TABernacle is a tool which presents Wikidata content in a tabular format. Here, the Item column shows a Wikidata identifier and its English label; The Label column displays labels for any of the target languages; and the Description column is the explanation text. Here, the Bangla fields (bn) are blank, the Hindi fields (hi) are incomplete, and the English fields (en) are used as starting points for curating the data in the other languages [57].

**Figure 5.**
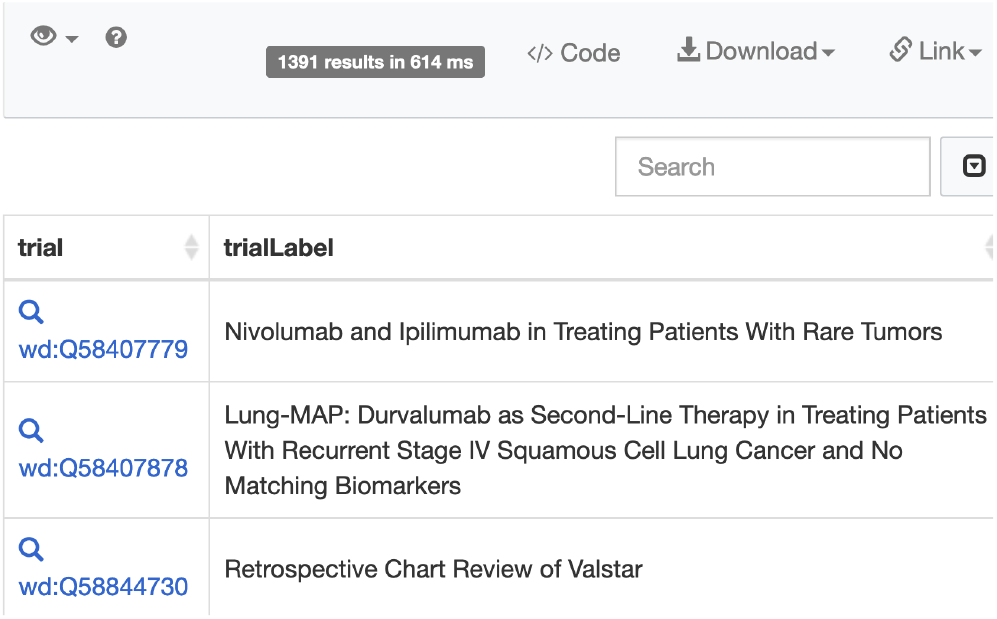
[Clinical Trials at Vanderbilt University](https://commons.wikimedia.org/wiki/File:Wikidata_Query_Service_-_WikiProject_Clinical_Trials_-_Clinical_trials_at_Vanderbilt_University.png). The Wikidata Query Service returns results for the query “[Clinical Trials at Vanderbilt University](https://www.wikidata.org/w/index.php?oldid=1585305957##Clinical_trials_at_Vanderbilt_University)” presents the Wikidata identifier and link to the full Wikidata record for each trial along with the trial’s name. The set of 1391 results are a profile of the research at this university.

The intent of providing profiles is to increase user engagement with clinical trials data by making it easier to access, share, republish, or discuss. Profiles can illustrate stories which users may wish to tell, like what sort of research happens at a university, the extent to which a timeline count of clinical trials by medical condition reflects interest in a field, or which organizations are involved most in research with certain characteristics. When WikiProject Clinical Trials provides a reproducible pathway for individuals and communities to subset and present representations of trial data, then the outcome of that pathway is a profile.

In some cases, we implemented standard profiles in Scholia, a Wikidata front end which assists with curation and profiling in Wikidata.[36] It is an online Wikidata tool that combines the results of multiple queries with some shared theme into web pages with multiple visualizations, and that has simple URLs for profiling various aspects of research based on the Wikidata identifier for the entity to be profiled.

Scholia users may be people who simply want easy access to existing content profiles related to clinical trials and do not want to understand much about Wikidata, or they may have deep understanding of Wikidata and want to use or contribute to such profiles, or they may find themselves somewhere in between.

### Other Wikidata activities

This project included other activities within the Wikidata platform which we summarize as communicating with the Wikidata community. Engagement with Wikidata requires engagement with an online community, and predicting precise outcomes from such human social interaction is not possible. Our advice to others who would consider replicating our process is to share enough documentation of your own activities to gain acceptance from the Wikidata community. This project’s primary channel of public communication was the WikiProject Clinical Trials project page, where reviewers can learn about the project, access its resources, and discuss issues.

Wikidata permits a variety of data remixing activities which are not formally documented, often without even informal documentation, and beyond the scope of the current project to document. One of the explanations for how the Wikimedia community promotes conformity is by maintaining a “community of practice”,[59], which means that proficiency in Wikimedia editing comes from practice and social interactions rather than following easily defined rules.[60] Beyond the technical engagement options, Wikidata encourages human-to-human conversation about the editorial process. Conversations may happen on documentation pages of data records and guidelines or in forums where people may discuss social, ethical, and technical issues related to activities in the platform. The Wikidata community expects that all people contributing data will respect the culture of its community, including by sharing quality content and avoiding editing in a way contrary to community norms.[12]

Figure 2 illustrates steps which may satisfy both content contributors and the regular Wikidata community. After importing a small bit of content to Wikidata, seek review of the shared content, and document activities in a WikiProject as a public forum. In turn, active members of a WikiProject will encourage more review, edit content, or do outreach to recruit other reviewers or expert contributions. When contributors come to an end point in a project, they should share the use case with a WikiProject, which again promotes transparency for review. Any Wikidata process which recruits more community review and consensus is less likely to cause a problem. Carelessness in Wikidata editing results in wastage of Wikidata volunteer time and resources, and may lead to community offense.

## Results

### WikiProject Clinical Trials as a resource portal

WikiProject Clinical Trials presents the features of a Wikidata WikiProject, including a “Models” page with a description and explanation of the data modeling for clinical trials and the “Queries” page with sample queries for accessing subsets of clinical trials data. Expected behavior of WikiProject Clinical Trials users includes querying and reviewing Wikidata clinical trial records, submitting additional edits, sharing profiles to encourage exploration, curation and discussion, and exporting data for reuse in other projects.

### Data integration and enrichment in Wikidata

The February 2022 snapshot of WikiProject Clinical Trials contains queries and results which measure the extent of content integration with Wikidata.[3] The “Count of clinical trials” query in our snapshot found 356,915 Wikidata records. The “Most common properties applied to clinical trials” query reports that 356,551 of those records are matched to a ClinicalTrials.gov ID, and number of records with “count of participants” and “study type” are similarly near complete, because these were easy to import without entity disambiguation or other curation. 320,021 records have “minimum age” listed and 177,418 have a “clinical trial phase”, which may indicate either that the original records lacked this information or that there was a difficulty in identifying structured data to import. Examples of more difficult properties to feature are “sponsor”, where there were 65,423 instances counted; and “funder”, where there were 133 instances. Challenges to importing sponsors were that Wikidata did not have records for all of the organizations in the original datasource, or when there were records, our process was not able to disambiguate the original prose to a Wikidata record. Funder data was not in the original datasource, and was our experiment to enrich the original content with public information we identified about a known funding organization. There are 17 properties applied in more than 50,000 instances and 56 properties applied to 10 trials or more. The ease with which each property value imports to Wikidata could be its own story and interpretation, but as this project is only presenting initial results, it is beyond the present scope to analyze all of these. The general rule was that when entity disambiguation was not required, then data import occurred more completely. Note also that the “Most common property” results count instances of property usage and not counts of clinical trials which have the properties, and that some properties often appear multiple times in a single trial record.

### Profiles for exploration and engagement

WikiProject Clinical Trials presents example profiles of clinical trials by medical condition, research intervention, research site, principal investigator, and funder. Users may view any of these profiles by running the query through the corresponding link in the WikiProject Clinical Trials “Query” tab. By modifying the identifiers used in query, users can profile any arbitrary topic of the same property class, such as exchanging one medical condition for another. Queries return results whenever data is available. As an example, Figure 6 displays a profile counting the appearance of research sites on trial records. Example further exploration based on this query could be the inverse - what trials a given site has hosted - or other details like what medical conditions or interventions were examined in trials at a given site.

**Figure 6.**
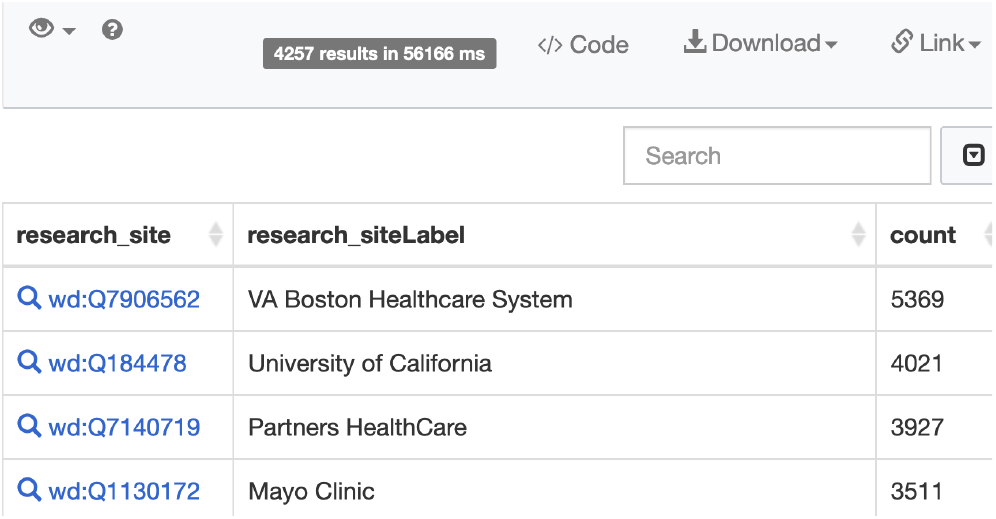
[Profile of Research sites](https://commons.wikimedia.org/wiki/File:Wikidata_Query_Service_-_WikiProject_Clinical_Trials_-_Research_sites.png). The Wikidata Query Service results for the the WikiProject Clinical Trials query “Research sites” counts Wikidata records which identify an organization or its subsidiaries as a research site in a clinical trial. The February 2022 snapshot shown here found 4257 different research sites, with “VA Healthcare System Boston” appearing research site in 5369 records.[3]

Any data imports in Wikidata interconnects with Wikidata’s other content holdings. The profile “Clinical trials for Zika fever” which is the model profile for “clinical trials by medical condition” lists those trials where the main subject (property P921) is Zika fever (item Q8071861). This medical condition was chosen because of development with a related project, WikiProject Zika Corpus, which profiled Zika virus in Wikidata [61] and developed norms in managing medical publications on Wikidata, thereby also paving the way for addressing the current COVID-19 pandemic in a systematic fashion [19]. ClinWiki contributed structured data and guidance to WikiProject Clinical Trials for organizing volunteers to apply topic tags of patient interest to clinical trial records. This organization also provided classification data to clinical trials which disambiguated unclear information in the original records, and which also demonstrated that a combination of expert review, community volunteering, and automation can enrich ClinicalTrials.gov data to identify medical conditions associated with trials. By showcasing WikiProject Zika Corpus and ClinWiki as projects with topic curation practices, WikiProject Clinical Trials sought to confirm norms in curation of such information in Wikidata.

### Language translation

We identified about 200 key terms related to clinical trials, matched those terms to corresponding Wikidata items, then organized language translation of those Wikidata items into Hindi, Bangla, and Swahili. One result of this activity is that we put these translated terms into Wikidata, where anyone can find, interact with, or reuse them. There are multiple dimensions of reuse of these translations, e.g. when anyone uses the Wikidata Query Service to seek content related to clinical trials, their query input and output can now come in these languages (cf. Figure 7).

**Figure 7.**
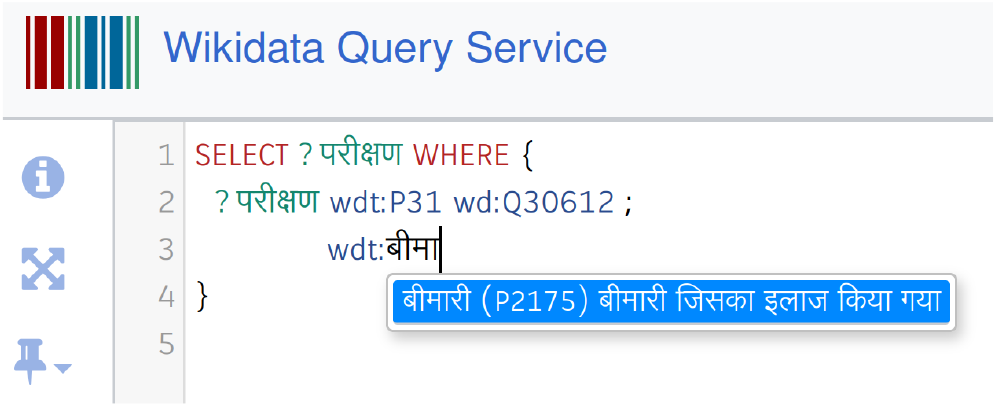
Autocompletion in the Wikidata Query Service in Hindi while building a query that uses the property P2175 (medical condition treated). When the user interface language is set to Hindi, the user can type the elements of their query in Hindi and get auto-suggestions that make use of the Wikidata labels and descriptions of the kind that were translated through this project. Labels for property P2175 are available in about 40 languages.

This project translated the items in the ClinicalTrials.gov publication, “Glossary of Common Site Terms”[58] and also the Wikidata property labels themselves which appear in Wikidata queries and the showcased query results. After the concepts of these terms were mapped to their equivalent Wikidata items, translation services were used to generate appropriate mappings of these concepts between English and the target languages. Additional review was sought for these potential translations, after which they were published in Wikidata for ongoing community review and engagement.

Outside of the project but complementary to the translation activity, the Wikidata community of editors has other translation projects in process, such as for the names of medical conditions and treatments. The “Curate” page of WikiProject Clinical Trials shares more about Wikimedia translation methods, including links to tools in the Wikimedia platform which support translation and pointers to Wikimedia community organizations which have a reputation for doing translation. Examining the tools or community activity can demonstrate the diversity of engagement with the Wikimedia translation.

## Discussion

This project did not produce a feature comparison of WikiProject Clinical Trials versus competing products and information sources. Outcomes which WikiProject Clinical Trials sought were an increase in accessibility and quality through its integration and curation in Wikidata. We demonstrated data import and the application of Wikidata browsing and curation processes, but comparing the various features of Wikidata with other products is challenging as each platform and dataset has its own strengths and weaknesses. An obvious question is what Wikidata can do that the ClinicalTrials.gov interface cannot. Answers include that Wikidata provides a different suite of tools for accessing and sorting the data, options to alter data, collaboration with an engaged user community, and a history of reuse by entities which collect data or operate search engines. Drawbacks to using Wikidata include the incompleteness of the data import, lack of trust in the editorial process, and new social complications from making data more accessible.

Wikimedia development proceeds stepwise. If Wikidata were to eventually have complete clinical trials data, then a possible driver toward this end could be groups of stake-holders who curated subsets of data which were interesting to them. For example, all of the clinical trials at a university could become structured when the university itself, students of the university, or some activist group curated those trials for their own purposes. Likewise a research network or patient advocacy group might curate all of the trials related to a medical condition or treatment. Individual researchers may want structured data for trials where they were contributors so that structured data circulates around the Internet to better showcase their accomplishments. The Wikidata community curates data for public benefit as a matter of ideology, and seeks to open whatever content it identifies as benefiting the most people. Whatever the way forward, WikiProject Clinical Trials has demonstrated methods for anyone to do such curation in Wikidata. Barriers to joining Wikidata are relatively low as compared to other projects, and data in Wikidata has reasonable potential to circulate around the Internet if anyone has increased visibility of content as their goal.

Querying Wikidata brings new perspectives to data, and seeing the data in new ways can make deficiencies in the original dataset more visible. Figure 8 shows a Scholia profile of “Recent clinical trials” where implausible start times of the year 2100 and 2050 are imported to Wikidata from the original records. The nature of the Wikimedia platform is to encourage users to flag and discuss odd results. There is currently no established communication channel back from Wikidata to other data sources if anyone edits content in Wikidata.

**Figure 8.**
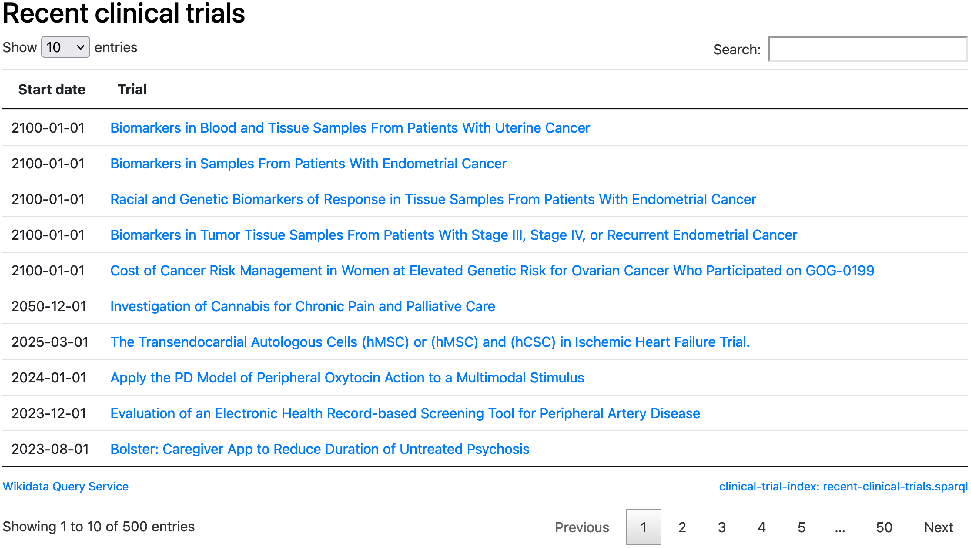
[Recent clinical trials - Scholia profile](https://commons.wikimedia.org/wiki/File:Scholia_-_Clinical_trials_overview_-_2021-12_-_Recent_clinical_trials_01.png) Scholia is a Wikidata front end which generates scholarly profiles. In this profile of “Recent clinical trials” it identifies trials registered to start in the year 2100, which must be an error. By querying trials and getting new perspectives, errors in the original records sometimes became visible.

Wikidata is an environment where social interactions include relationships between humans and machines; experts and amateurs with respect to subject matter (including patients, researchers, clinicians); and people from different cultural backgrounds, both within and across countries. Some of these relationships surfaced during the development of the comparable WikiProject COVID-19. One difference between the projects, though not necessarily the most significant, is that the community at WikiProject COVID-19 has its own collection of queries and datasets which raise different issues that those of this project.[19]. Lowering access barriers to clinical trials data could have unexpected social and ethical consequences. The original dataset at ClinicalTrials.gov was already available to the public as a service of the United States government. Our intent in setting up this project was to make this data yet more accessible. If our approach would be successful to the limits of our imagination, then clinical trial data would become so accessible that any typical person could ask and receive answers to many routine questions pertaining to clinical trials. Example questions that people might have are, “What clinical trials are happening in my community?” or “Which organizations do clinical trials for my condition?” or combinations thereof, such as “Which organizations near me do clinical trials for my condition and are currently recruiting participants with my demographics?” Even if Wikidata is not the platform to eventually provide universal access to such questions, then eventually, the data will be open enough and tools will be accessible enough that somehow there will be increased access to this information. WikiProject Clinical Trials and the Wikidata platform can be a place for live and public community examination of sensitive social issues, inequalities in the distribution of clinical research, mismatches between community expectations and research priorities, or whatever other social issues can arise when transparency increases.

The approach which WikiProject Clinical Trials applied to integrate ClinicalTrials.gov into Wikidata could apply to other clinical trials registries. The WHO’s International Clinical Trials Registry Platform has been critiqued for lacking metadata to facilitate search,[62] and curating metadata in Wikidata could address difficulties in accessing records which that platform indexes. While ClinicalTrials.gov is available only in English, there are registries with primary languages other than English and limited information transfer across languages.[63] Wikidata could be a platform for translation and data sharing across these registries and similar resources. Similarly, many clinical trials include research sites in multiple countries. For any set of trials which recruit non-English speakers, WikiProject Clinical Trials presents public options for contributing translations. If it ever happens that an activist group of community stakeholders in clinical research made a request to the pharmaceutical industry that clinical trial metadata should be available in the language of research participants, then they could point to WikiProject Clinical Trials and the Wikidata platform as an outlet for hosting, maintaining, and distributing that content.

## Conclusions

WikiProject Clinical Trials imported ClinicalTrials.gov data to Wikidata, developed a process for generating browser-based metadata profiles of sets of clinical trials, and demonstrated activities which invite public engagement with the data to explore, refine, remix and reuse it. The data import was less complete where the original dataset was ambiguous. Wikidata curation included entity disambiguation and language translation. Users who are interested in further such data curation may contribute to Wikidata themselves, or export Wikidata content for reuse. Both contributions and reuse happen in multiple ways and at scale.

The documentation presented here of WikiProject Clinical Trials may be insightful as a model for how researchers can use Wikidata as a platform to recruit community engagement with datasets. Advantages of using Wikidata as a virtual environment include that it has an existing user community and tools for data curation. The Wikimedia environment is public and shared, so data imports trigger social reactions. While community rules are not always documented in detail, friendly collaboration, transparent communication, and respect for the time and labor of volunteers fulfills many expectations.

## Data Availability

All data produced are available online as referenced and linked in manuscript.

https://www.wikidata.org/wiki/Wikidata:WikiProject_Clinical_Trials

https://doi.org/10.5281/zenodo.6317047

## Data and software availability

Table 4, “Software used by WikiProject Clinical Trials”, links to either a running instance of tools or the corresponding code repository, and provides both Research Resource and Wikidata identifiers.

Table 5, “Data used by WikiProject Clinical Trials”, links to project pages managing resources and again provides identifiers.

**Table 5.** Data used by WikiProject Clinical Trials. The dataset column is the name and link, while RRID and Wikidata items are identifiers assigned by SciCrunch and Wikidata.

| Dataset | RRID | Wikidata item |
| --- | --- | --- |
| Aggregate Analysis of ClinicalTrials.gov | SCR_021247 | Q76693569 |
| ClinicalTrials.gov | SCR_021296 | Q5133746 |
| ClinWiki | SCR_021297 | Q96777166 |
| ClinicalTrials.gov Glossary |  | Q106891230 |
| WikiProject Clinical Trials snapshot February 2022 |  | Q111145762 |
| WikiProject Zika Corpus | SCR_022079 | Q54439832 |

## Consent

No consent was required for research or publication. This project used data that was public from creation, and that contained no elements that anyone identified as private.

## Author contributions

Credit to each author is as follows:

- LR - conceptualization, methodology, funding acquisition, supervision, writing (original draft preparation)
- ST - methodology, software, investigation, writing (review and editing)
- WH - data curation, supervision, resources, software
- AW - data curation
- JK - data curation, methodology
- SB - methodology, software, visualization, formal analysis
- CA - conceptualization, data curation, visualization, validation, methodology
- PW - writing (original draft preparation), resources, supervision
- CK - writing (review and editing)
- DM - conceptualization, methodology, funding acquisition, visualization, investigation, validation, writing (review and editing)

## Acknowledgements

WikiProject Clinical Trials is a Wikimedia project, and as such, is built upon the shared public media commons. Thanks particularly to the following:

- WikiCite, Wikidata, and Wikipedia communities
- the Aggregate Analysis of ClinicalTrials.gov (AACT) project team for sharing this project’s primary imported dataset
- Librarians at Vanderbilt University for sharing Wikidata’s most complete university research corpus
- ClinWiki community of volunteer editors for sharing disambiguated clinical trial record data
- The Patient-Centered Outcomes Research Institute for referrals to published material about their projects
- Scholia developers, including Finn Årup Nielsen, Egon Willighagen, and Carlin MacKenzie, for developing Wikidata-based profiling
- Tanvir Rahman and Wikimedia Bangladesh for consultation on translation

## References

[1] Andra Waagmeester, Gregory Stupp, Sebastian Burgstaller-Muehlbacher, Benjamin M. Good, Malachi Griffith, Obi Griffith, Kristina Hanspers, Henning Hermjakob, Toby Hudson, Kevin Hybiske, Sarah M Keating, Magnus Manske, Michael Mayers, Daniel Mietchen, Elvira Mitraka, Alexander R. Pico, Timothy Elliott Putman, Anders Riutta, Núria Queralt Rosinach, Lynn Schriml, Thomas Shafee, Denise Slenter, Ralf Stephan, Katherine Thornton, Ginger Tsueng, Roger Tu, Sabah Ul-Hasan, Egon Willighagen, Chunlei Wu, and Andrew I. Su. Wikidata as a knowledge graph for the life sciences. eLife, 9, mar 17 2020.

[2] Wikimedia community. Wikidata:WikiProject Clinical Trials, 2022. URL https://www.wikidata.org/wiki/Wikidata:WikiProject_Clinical_Trials.

[3] Lane Rasberry and Daniel Mietchen. WikiProject Clinical Trials snapshot February 2022, February 2022. URL https://doi.org/10.5281/zenodo.6317047.

[4] Tony Tse, Rebecca J Williams, and Deborah A Zarin. Reporting “basic results” in https://clinicaltrials.gov. Chest, 136(1): 295–303, 2009.

[5] Deborah A Zarin, Tony Tse, Rebecca J Williams, Robert M Califf, and Nicholas C Ide. The ClinicalTrials.gov results database–update and key issues. The New England Journal of Medicine, 364(9):852–860, mar 1 2011.

[6] Danny T Y Wu, David A Hanauer, Qiaozhu Mei, Patricia M Clark, Lawrence C An, Joshua Proulx, Qing T Zeng, V G Vinod Vydiswaran, Kevyn Collins-Thompson, and Kai Zheng. Assessing the readability of ClinicalTrials.gov. Journal of the American Medical Informatics Association, 23(2): 269–275, aug 11 2015.

[7] Asba Tasneem, Laura Aberle, Hari Ananth, Swati Chakraborty, Karen Chiswell, Brian J. McCourt, and Ricardo Pietrobon. The Database for Aggregate Analysis of ClinicalTrials.gov (AACT) and Subsequent Regrouping by Clinical Specialty. PLoS ONE, 7(3):e33677, March 2012. ISSN 1932-6203. doi: 10.1371/journal.pone.0033677. URL https://dx.plos.org/10.1371/journal.pone.0033677.

[8] Laura Miron, Rafael S Gonçalves, and Mark A. Musen. Obstacles to the reuse of study metadata in ClinicalTrials.gov. Scientific Data, 7(1):443, ec 18 2020.

[9] Raghu Garud, Sanjay Jain, and Philipp Tuertscher. Incomplete by design and designing for incompleteness. Organization studies, 29(3):351–371, 2008.

[10] Chitu Okoli, Mohamad Mehdi, Mostafa Mesgari, Finn Årup Nielsen, and Arto Lanamäki. Wikipedia in the eyes of its beholders: A systematic review of scholarly research on wikipedia readers and readership. Journal of the Association for Information Science and Technology, 65(12):2381–2403, 2014.

[11] Mostafa Mesgari, Chitu Okoli, Mohamad Mehdi, Finn Årup Nielsen, and Arto Lanamäki. “the sum of all human knowledge”: A systematic review of scholarly research on the content of wikipedia. Journal of the Association for Information Science and Technology, 66(2): 219–245, 2015.

[12] Dariusz Jemielniak. Common knowledge? Stanford University Press, 2014. ISBN 978-0804789448.

[13] James M Heilman and Andrew G West. Wikipedia and medicine: quantifying readership, editors, and the significance of natural language. Journal of medical Internet research, 17(3):e4069, 2015.

[14] Denny Vrandečić and Markus Krötzsch. Wikidata: a free collaborative knowledgebase. 57:78–85, 10 2014.

[15] Alessandro Piscopo, Christopher Phethean, and Elena Simperl. Wikidatians are born: paths to full participation in a collaborative structured knowledge base. 1 2017.

[16] Alexander Pfundner, Tobias Schönberg, John Horn, Richard D Boyce, and Matthias Samwald. Utilizing the Wikidata system to improve the quality of medical content in Wikipedia in diverse languages: a pilot study. Journal of Medical Internet Research, 17:e110, may 5 2015.

[17] Houcemeddine Turki, Thomas Shafee, Mohamed Ali Hadj Taieb, Mohamed Ben Aouicha, Denny Vrandečić, Diptanshu Das, and Helmi Hamdi. Wikidata: A large-scale collaborative ontological medical database. 99:103292, sep 23 2019.

[18] Denise A Smith. Situating Wikipedia as a health information resource in various contexts: A scoping review. PloS one, 15(2):e0228786, 2020.

[19] Houcemeddine Turki, Mohamed Ali Hadj Taieb, Thomas Shafee, Tiago Lubiana, Dariusz Jemielniak, Mohamed Ben Aouicha, Jose Emilio Labra Gayo, Eric A. Youngstrom, Mus’ab Banat, Diptanshu Das, Daniel Mietchen, and WikiProject COVID-19. Representing COVID-19 information in collaborative knowledge graphs: The case of Wikidata. Semantic Web: Interoperability, Usability, Applicability, pages 1–32, sep 28 2021. URL https://doi.org/10.3233/SW-210444.

[20] Lucie-Aimée Kaffee, Alessandro Piscopo, Pavlos Vougiouklis, Elena Simperl, Leslie Carr, and Lydia Pintscher. A Glimpse into Babel: An Analysis of Multilinguality in Wikidata. 8 2017.

[21] Lucie-Aimée Kaffee, Kemele M. Endris, and Elena Simperl. When Humans and Machines Collaborate: Cross-lingual Label Editing in Wikidata. In Proceedings of the 15th International Symposium on Open Collaboration, 8 2019.

[22] Daniel Mietchen, Gregor Hagedorn, Egon Willighagen, Mariano Rico, Asunción Gómez-Pérez, Eduard Aibar, Karima Rafes, Cécile Germain, Alastair Dunning, Lydia Pintscher, and Daniel Kinzler. Enabling Open Science: Wikidata for Research (Wiki4R). 1:e7573, ec 22 2015.

[23] Sebastian Burgstaller-Muehlbacher, Andra Waagmeester, Elvira Mitraka, Julia Turner, Tim Putman, Justin Leong, Chinmay Naik, Paul Pavlidis, Lynn Schriml, Benjamin M Good, et al. Wikidata as a semantic framework for the gene wiki initiative. Database, 2016, 2016.

[24] Tim E. Putman, Sebastien Lelong, Sebastian Burgstaller-Muehlbacher, Andra Waagmeester, Colin Diesh, Nathan Dunn, Monica Munoz-Torres, Gregory S. Stupp, Chunlei Wu, Andrew I. Su, and Benjamin M. Good. WikiGenomes: an open web application for community consumption and curation of gene annotation data in Wikidata. Database, 2017, 03 2017. ISSN 1758-0463. doi: 10.1093/database/bax025. URL https://doi.org/10.1093/database/bax025.bax025.

[25] Mariam Farda-Sarbas and Claudia Müller-Birn. Wikidata from a Research Perspective – A Systematic Mapping Study of Wikidata. aug 29 2019.

[26] Julie McDonough Dolmaya. Expanding the sum of all human knowledge: Wikipedia, translation and linguistic justice. The Translator, 23(2):143–157, 2017.

[27] Wikimedia community. Wikimedia projects, 2022. URL https://meta.wikimedia.org/wiki/Wikimedia_ projects.

[28] Mohamad Mehdi, Chitu Okoli, Mostafa Mesgari, Finn Årup Nielsen, and Arto Lanamäki. Excavating the mother lode of human-generated text: A systematic review of research that uses the wikipedia corpus. Information Processing & Management, 53(2):505–529, 2017.

[29] Denny Vrandečić and Markus Krötzsch. Wikidata: a free collaborative knowledgebase. Communications of the ACM, 57(10):78–85, September 2014. ISSN 0001-0782, 1557-7317. doi: 10.1145/2629489. URL https://dl.acm.org/doi/10.1145/2629489.

[30] Stanislav Malyshev, Markus Krötzsch, Larry González, Julius Gonsior, and Adrian Bielefeldt. Getting the most out of wikidata: Semantic technology usage in wikipedia’s knowledge graph. In Denny Vrandečić, Kalina Bontcheva, Mari Carmen Suárez-Figueroa, Valentina Presutti, Irene Celino, Marta Sabou, Lucie-Aimée Kaffee, and Elena Simperl, editors, The Semantic Web – ISWC 2018, pages 376–394, Cham, 2018. Springer International Publishing. doi: 10.1007/978-3-030-00668-6_23.

[31] Stacy Allison-Cassin, Alison Armstrong, Phoebe Ayers, Tom Cramer, Mark Custer, Mairelys Lemus-Rojas, Sally McCallum, Merrilee Proffitt, Mark Puente, Judy Ruttenberg, et al. ARL white paper on Wikidata: Opportunities and recommendations. 2019.

[32] Anne Britton. Wikidata as a Tool for Mapping Investment in Open Infrastructure: An Exploratory Study. Dec 17 2021.

[33] ClinWiki. About us, 2022. URL https://www.clinwiki.org/about-us.

[34] Vanderbilt University Research News. Librarians work to broaden vanderbilt’s research reputation with wikidata tools, 08 2020. URL https://news.vanderbilt.edu/2020/08/10/librarians-work-to-broaden-vanderbilts-research-reputation-with-wikidata-tools/.

[35] Joseph V Selby and Steven H Lipstein. Pcori at 3 years–progress, lessons, and plans. The New England Journal of Medicine, 370(7):592–595, feb 1 2014.

[36] Finn Årup Nielsen, Daniel Mietchen, and Egon Willighagen. Scholia, scientometrics and wikidata. In European Semantic Web Conference, pages 237–259. Springer, 2017.

[37] Mairelys Lemus-Rojas. Exploring the potential of wikidata & scholia to generate scholarly profiles at iupui. 2018.

[38] Wikimedia community. WikiProject Medicine, 2022. URL https://en.wikipedia.org/wiki/Wikipedia:WikiProject_Medicine.

[39] Richard James. Wikiproject Medicine: Creating Credibility in Consumer Health. Journal of Hospital Librarianship, 16 (4):344–351, 10 2016.

[40] Lane Rasberry and Daniel Mietchen. Fair and open multilingual clinical trials in Wikidata and Wikipedia. Research Ideas and Outcomes, 7, 25 March 2021. doi: 10.3897/rio.7.e66490.

[41] Wikidata community. Wikidata help:contents, 2022. URL https://www.wikidata.org/wiki/Help:Contents.

[42] Wikimedia community. Wikidata:WikiProject Clinical Trials, 2022. URL https://www.wikidata.org/wiki/Wikidata:WikiProject_Clinical_Trials/Model.

[43] Andra Waagmeester, Egon L. Willighagen, Andrew I. Su, Martina Kutmon, Jose Emilio Labra Gayo, Daniel Fernández-Álvarez, Quentin Groom, Peter J. Schaap, Lisa M. Verhagen, and Jasper J. Koehorst. A protocol for adding knowledge to Wikidata: aligning resources on human coronaviruses. 19:12, jan 22 2021.

[44] Eva Seidlmayer, Jakob Voß, Tatyana Melnychuk, Lukas Galke, Klaus Tochtermann, Carsten Schultz, and Konrad U Förstner. Orcid for wikidata. data enrichment for scientometric applications. In 1st Wikidata Workshop (Wikidata 2020). CEUR Workshop Proceedings, 2020.

[45] Wikidata community. Wikidata:property proposal, 2022. URL https://www.wikidata.org/wiki/Wikidata:Property_proposal.

[46] Wikimedia community. Wikidata:copyright, 2022. URL https://www.wikidata.org/wiki/Wikidata:Copyright.

[47] Creative Commons. Cc0 1.0 universal (cc0 1.0) public domain dedication, 2022. URL https://creativecommons.org/publicdomain/zero/1.0/.

[48] Clinical Trials Transformation Initiative. Wikidata:wikiproject clinical trials, 2022. URL http://www.ctti-clinicaltrials.org.

[49] Andrea Forte, Vanesa Larco, and Amy Bruckman. Decentralization in wikipedia governance. Journal of Management Information Systems, 26(1):49–72, 2009.

[50] Lei Zheng, Christopher M Albano, Neev M Vora, Feng Mai, and Jeffrey V Nickerson. The roles bots play in wikipedia. Proceedings of the ACM on Human-Computer Interaction, 3 (CSCW):1–20, 2019.

[51] Adrian Bielefeldt, Julius Gonsior, and Markus Krötzsch. Practical linked data access via sparql: the case of wikidata. In LDOW@ WWW, 2018.

[52] Wikimedia community. Wikidata:WikiProject Clinical Trials, 2022. URL https://www.wikidata.org/wiki/Wikidata:WikiProject_Clinical_Trials/Queries.

[53] Alessandro Piscopo and Elena Simperl. What we talk about when we talk about wikidata quality: a literature survey. In Proceedings of the 15th International Symposium on Open Collaboration, pages 1–11, Skövde Sweden, August 2019. ACM. ISBN 978-1-4503-6319-8. doi: 10.1145/3306446.3340822. URL https://dl.acm.org/doi/10.1145/3306446.3340822.

[54] Wikimedia community. Wikidata:WikiProject Clinical Trials, 2022. URL https://www.wikidata.org/wiki/Wikidata:WikiProject_Clinical_Trials/Curate.

[55] Oliver Ferschke, Torsten Zesch, and Iryna Gurevych. Wikipedia revision toolkit: Efficiently accessing wikipedia’s edit history. In Proceedings of the ACL-HLT 2011 System Demonstrations, pages 97–102, 2011.

[56] Liam Wyatt, Phoebe Ayers, Merrilee Proffitt, Daniel Mietchen, Dario Taraborelli, Alex Stinson, Amanda Bittaker, Jonathan Curiel, Janice Tud, and Caitlin Virtue. WikiCite 2020-2021: Citations for the sum of all human knowledge. Technical report, Zenodo, October 2021. URL https://zenodo.org/record/5363757.

[57] Daniel Mietchen. 1000 Disease Ontology terms and their Wikidata mappings to 17 mostly Indian languages and English. Feb 13 2020. doi: 10.5281/zenodo.3666921. URL https://doi.org/10.5281/zenodo.3666921.

[58] http://ClinicalTrials.gov. Glossary of common site terms, 2021. URL https://clinicaltrials.gov/ct2/about-studies/glossary.

[59] Dan O’Sullivan. Wikipedia: a new community of practice? Routledge, 2016.

[60] Jean Lave and Etienne Wenger. Situated learning: Legitimate peripheral participation. Cambridge university press, 1991.

[61] Sean Ekins, Daniel Mietchen, Megan Coffee, Thomas P Stratton, Joel S Freundlich, Lucio Freitas-Junior, Eugene Muratov, Jair Siqueira-Neto, Antony J Williams, and Carolina Andrade. Open drug discovery for the Zika virus. F1000Research, 5(150):150, 2016.

[62] Julie M Glanville, Steven Duffy, Rachael McCool, and Danielle Varley. Searching clinicaltrials. gov and the international clinical trials registry platform to inform systematic reviews: what are the optimal search approaches? Journal of the Medical Library Association: JMLA, 102(3): 177, 2014.

[63] Daisuke Ogino, Kunihiko Takahashi, and Hajime Sato. Characteristics of clinical trial websites: information distribution between ClinicalTrials.gov and 13 primary registries in the WHO registry network. Trials, 15(1):1–8, 2014.

